## Appendices for "Communicating personalised risks from COVID-19: guidelines from an empirical study"

### Appendix 1

Characteristics of participants in the qualitative interviews

| Phase | Number of participants | Ages | Genders | Ethnicities | Experience with COVID |
| --- | --- | --- | --- | --- | --- |
| Discovery - public | 6 | 22-73 | 2 male, 4 female | 5 white, 1 BAME | 1 had a relative die from COVID-19, and knew others who had it (P4) |
| Alpha round 1 | 6 | 34-76 | 4 male, 2 female | 5 white, 1 BAME | 1 knew someone who had it (P10) |
| Alpha round 2  (a & b) | 8 | 19-87 | 4 male, 4 female | 7 white, 1 BAME | 1 had symptoms, assumed COVID-19 (P19) 5 knew people who had it (P13; P16; P17; P19; P20)  3 knew people who died from it (P14; P18; P20) |
| Alpha round 3 | 8 | 23-71 | 4 male, 4 female | 7 white, 1 BAME | 1 has symptoms, assumes COVID-19, was awaiting test results (P23)  3 knew people who had it (P24; P27; P28) 2 knew people who died from it (P26; P22) |
| Discovery - primary care physicians | 7 | 35-65 | 2 male, 5 female | 5 white, 2 BAME | 6 out of 7 GPs had patients who were suspected of having it/did have it (1 GP wasn't practising throughout the pandemic). They came across it frequently during the peak, but less so by the time of the interviews. |

### Appendix 2

Participants’ numerical interpretations of ‘high’ and ‘low’ risk of dying from COVID-19 during interviews

| Participant number | ‘Low risk’ estimate | ‘High risk’ estimate |
| --- | --- | --- |
| 11 | 1/10 | 10/10 |
| 7 | 1/10 | 8/10 |
| 16 | - | 90% |
| 13 | <50% | >90% |
| 20 | 1% | 50% |
| 18 |  | 100% |
| 21 | <20% | - |
| 22 | <3/10 | 7/10 |
| 23 | 2/10 | 8/10 |
| 24 | <1% | 70% |
| 25 | <40% | >70% |

Key questions asked in quantitative surveys

### Appendix 3

Summary of key questions asked in each quantitative survey (grouped by theme)

| Theme | Survey | Questions asked | Answer options |
| --- | --- | --- | --- |
| What information the audience want | 1, 2, 3 | I would like to know now what my personal risk of dying from COVID-19 would be if I were to catch the virus  I think that people are entitled to know now what their personal risk of dying from COVID-19 would be if they were to catch it  I would not like my employer to know my personal risk of dying from COVID-19  I feel that I have enough information already about my personal risks from COVID-19  I would like to know by how much each personal behavioural change (e.g. wearing face masks, washing hands) reduces my personal risk of catching the virus  I would not trust any information about my personal risk of dying from COVID-19 as I don’t believe enough is known about it | 7-point Likert scales: Completely disagree - Completely agree |
|  | 3 | Which of the following pieces of information would you most want to know about your risk, if it were available? Please drag and drop into the order you feel puts the most important at the top: The risk of you dying of COVID-19 if you caught it; The risk of you catching COVID-19 given where you live and how many other cases have been reported nearby (will vary day-to-day a bit like a pollen count); The risk of you being hospitalised by COVID-19 if you caught it; Whether you might suffer long-term complications from COVID-19 if you caught it | Rank order |
|  |  | Is there any other information about your risk from COVID-19 that you would like to know that we haven't mentioned? | Free text |
|  | 3 | Please drag and drop the following into the order you feel puts the most important at the top: The person's risk score: the chances of that person dying if they caught COVID-19; Information about what data was used to calculate that risk score (eg. whether it was data from the UK, how much data the researchers had);  Information about who developed the maths behind the calculation;  Information about the certainty and precision of the risk score: how much lower or higher the actual risk for that person could be; Context about that risk score: how it rates compared to a similar person at different ages (eg. 'someone like you but aged 20'; 'someone like you but aged 60');  Context about that risk score: where that person's risk lies in comparison to everyone else in the UK (eg. a graph of the risks of everyone in the UK with an arrow showing 'you are here' to make it clear how many people have a higher risk than that person, and how many a lower risk); Link to where people can get more information about things they can do to reduce their chances of catching COVID-19 (eg. hand washing, social distancing, wearing face masks etc); Information reminding people that as well as the risk to themselves, they also pose a risk to others (in case they catch COVID-19 and pass it on); Details about how the risk score was calculated (eg. what factors makes someone's risk higher or lower, and how important each of those factors are); Information about thing they can do to reduce their chances of dying from COVID-19 if they caught it (such as losing weight) | Rank order |
|  | 4 | “A tool like this can be designed in many different ways, which give people a different impression of the numbers. The tool can be made as neutral as possible, to try to just inform people and let them make up their own mind about the risk to them and how they should behave. Or the differences between numbers can be made very obvious, making the risk look larger or smaller. This would be more persuasive and make people more likely to change their behaviour. How do you think a national tool should be designed?”  (For those who marked 4-7 on question above):  “In which direction do you think the tool should try to persuade people?” | 7-point Likert scale: As neutral as possible: just inform people - As persuasive as possible: change people's behaviour  3 options:  It should try to reassure people by showing the risk is generally low; It should try to make people be more cautious by showing that even if the risk is low to them, they can spread it to others; It should try to be persuasive to different people in different ways (reassuring some and making others more cautious) |
| Perception of the risk of COVID-19/risks as percentages and frequencies | 2  Exp 2.1 | 4 x 2 experimental design:  “Imagine that you were told that if you caught COVID-19, your risk of dying of it were: [randomised within subjects: 0.1%, 1%, 2%, 12%, 20%; OR (between subjects): 1 in 1000, 10 in 1000, 20 in 1000, 120 in 1000, 200 in 1000] How would you classify that risk in your mind?”  “If [name] catches COVID-19, [name]’s risk of dying is: [randomised within subjects: 0.1%, 1%, 2%, 12%, 20%; OR (between subjects): 1 in 1000, 10 in 1000, 20 in 1000, 120 in 1000, 200 in 1000] How would you classify that risk in your mind? For comparison: The average 85 year old man with no health problems has a risk of 12%. The average 75 year old man with no health problems has a risk of 1.7%. The average man under 55 with no health problems has a risk of below 0.5%”  “If [name] catches COVID-19, [name]'s risk of dying is: [randomised within subjects: 0.1%, 1%, 2%, 12%, 20%; OR (between subjects): 1 in 1000, 10 in 1000, 20 in 1000, 120 in 1000, 200 in 1000] How would you classify that risk in your mind? For context: [Within subjects chosen to match the risk level: They are a white man aged 30 with no underlying health conditions; They are a mixed race man aged 30 with two underlying health issues; They are a white woman aged 40 with a high BMI and undergoing cancer treatment; They are a black woman aged 75 and with certain underlying health issues; They are an Asian man aged 85 with a heart condition and diabetes]”  “If [name] catches COVID-19, [name]'s risk of dying is: [randomised within subjects: 0.1%, 1%, 2%, 12%, 20%; OR (between subjects): 1 in 1000, 10 in 1000, 20 in 1000, 120 in 1000, 200 in 1000] How would you classify that risk in your mind? For context: [Within subjects chosen to contrast with the risk level: They are a white man aged 30 with no underlying health conditions; They are a mixed race man aged 30 with two underlying health issues; They are a white woman aged 40 with a high BMI and undergoing cancer treatment; They are a black woman aged 75 and with certain underlying health issues; They are an Asian man aged 85 with a heart condition and diabetes]” | slider with no numerical cues: ‘very low risk’ and ‘very high risk’ as the end points |
|  | 4  Exp 4.1 | Randomised within subjects:  Alex is a black woman aged 75 and with some underlying health issues/ Sam is an Asian man aged 85 with a heart condition and diabetes/ Jo is a white woman aged 40 with a high BMI and undergoing cancer treatment/ Ali is a mixed race man aged 30 with two underlying health issues/ Mel is a white man aged 30 with no underlying health conditions  If [*he/she]* caught COVID-19, what would you estimate were her chances of dying of it? | 3 arm experimental design:  “Type a percentage, without the percentage sign”  “Type the number of people out of 100 exactly like [*name*] you would expect to die if they all caught it.”  “Type the number of people out of 1000 exactly like [*name*] you would expect to die if they all caught it.” |
|  | 4  Exp 4.2 | 5 arm experimental design. 4 arms:  Imagine that you were told that if you caught COVID-19, your risk of dying of it would be: [randomised within subjects: 0.1%, 1%, 2%, 12%, 20%; OR (between subjects): 0.1 in 100, 1 in 100, 2 in 100, 12 in 100, 20 in 100; OR (between subjects): 1 in 1000, 10 in 1000, 20 in 1000, 120 in 1000, 200 in 1000; OR (between subjects): 1 in 1000, 1 in 100, 1 in 50, 1 in 8, 1 in 5] How would you classify that risk in your mind? | 4 arms:  slider with no numerical cues: ‘very low risk’ and ‘very high risk’ as the end points  5th arm:  given percentages but responded on an 11-point Likert scale: Very low risk - Very high risk (no mid-points labelled) |
| Effects of log versus linear scale | 3 | 2 x 2 x 2 experiment (log vs linear scale; result as frequency vs percentage; risk level high or low)  How well did you **understand** the information in the mock-up?  How **clear** is the information in the mock-up?  If the person who got this result caught COVID-19, how likely do you think it is that they would die as a result?  How would you describe the risk of this person dying from COVID-19 if they caught it?  If this result applied to you, how worried would you be?  Thinking just about the risk result given for the individual: the chances that they would die of COVID-19 if they caught it:  To what extent do you think that this number is certain or uncertain?  To what extent do you think this number is:  Accurate  Reliable  Trustworthy  To what extent do you think that the people responsible for producing this number are trustworthy?  (The below questions randomised presentation)  If this result applied to me, I would not be worried about catching COVID-19  I would NOT like to see information like this about my own risks from COVID-19  If this result applied to me, I would likely be anxious and it might affect my mental health  If this result applied to me, I would likely change my behaviour to be less cautious of catching the virus  If this result applied to me, I would do everything I could to avoid catching the virus  I would not like my employer to know a result like this about my personal risk from COVID-19  If this result applied to me, I would be happy to go to a crowded social event | 7-point Likert scale: Not at all – completely  Above 2 combined into an index measure of comprehension (r=0.82)  7-point Likert scale: Very unlikely; Unlikely; Somewhat unlikely; neither likely nor unlikely; Somewhat likely; Likely; Very likely  slider with no numerical cues: ‘very low risk’ and ‘very high risk’ as the end points  Above 2 combined into an index measure of cognitive risk perception (r=0.73)  7-point Likert scale: Not at all worried - Very worried  7-point Likert scale: Very certain; Certain; Somewhat certain; neither certain nor uncertain; Somewhat uncertain; Uncertain; Very uncertain  7-point Likert scale: Not at all - Very    7-point Likert scale: Not at all trustworthy - Very trustworthy  7-point Likert scales: Completely disagree - Completely agree |
| Positive versus negative framing/Use of a visual scale/Addition of risk comparators for context | 2  Exp 2.2 | 3 arm experimental design  (Scale with no comparators; scale with age comparators; scale with graphic to show distribution of risk in population)  How well did you **understand** the information in the mock-up?  How **clear** is the information in the mock-up?  If the person who got this result caught COVID-19, how likely do you think it is that they would die as a result?    If this result applied to you, how worried would you be?  How would you describe the risk of this person dying from COVID-19 if they caught it?  (The below questions randomised presentation)  If this result applied to me, I would not be worried about catching COVID-19  I would NOT like to see information like this about my own risks from COVID-19  If this result applied to me, I would likely be anxious and it might affect my mental health  If this result applied to me, I would likely change my behaviour to be less cautious of catching the virus  If this result applied to me, I would do everything I could to avoid catching the virus  I would not like my employer to know a result like this about my personal risk from COVID-19  If this result applied to me, I would be happy to go to a crowded social event | 7-point Likert scale: Not at all - completely    7-point Likert scale: Very unlikely; Unlikely; Somewhat unlikely; neither likely nor unlikely; Somewhat likely; Likely; Very likely  7-point Likert scale: Not at all worried - Very worried  slider with no numerical cues: ‘very low risk’ and ‘very high risk’ as the end points  7-point Likert scales: Completely disagree - Completely agree |
|  | 4  Exp 4.3 | 5 x 4 between-subjects experimental design  (Formats:  -positive framing, visual scale, no comparators;  -negative framing, visual scale, no comparators;  -negative framing, visual scale, flu risk as comparator;  -negative framing, visual scale, age risks as comparators;  - negative framing, text only, age risks as comparators  Risk level: 0.01%, 0.1%, 2%, 20%)  All the questions in the above experiment plus:  Communication efficacy scale (based on [Scheuner, 2013](https://doi.org/10.1038/gim.2012.151)):  How satisfied are you with the general format (look and feel) of the mock-up?  How satisfied are you with the amount of information in the mock-up?  How satisfied are you with the organization of the information in the mock-up?  How easy is it to find the person's risk result in the mock-up?  How easy is it to find information in the mock-up that helps you decide what to do?  How easy is it to understand the language used in the mock-up?  How easy is it to understand the result presented in the mock-up?  How easy is it to understand what the result in the mock-up actually means?  How effectively does the mock-up communicate the person's risk?  How effectively does the mock-up communicate what this result means?  How effectively does the mock-up communicate the person's options having received this risk result?  How effectively does the mock-up communicate the availability of further information for the person receiving it?  How effectively does the mock-up communicate the limitations of the result?  Actionability scale [(slightly modified from Recchia, 2020](https://doi.org/10.1038/s41436-019-0649-0)):  How clear are you about what actions you could take if you had received this result in real life? Do you feel you would have the necessary information to decide what actions to take if you had received this result in real life? How certain are you about what you would do next if you had received this result in real life? Do you feel you would have the necessary professional support to decide what to do next if you had received this result in real life? How ready would you feel to take any next steps if you had received this result in real life?  Approximately what percentage of people who got a result like this would you expect to survive COVID-19 if they caught it? Please enter a number between 0 and 100 (decimal points are fine).  Out of 10,000 people who got a result like this, how many would you expect to survive COVID-19 if they caught it?  If this result applied to me I'd be more concerned about my risk from COVID-19 than from seasonal 'flu  I think the tool should just tell people whether their risk is 'high' or 'low'  “How worried would you feel doing each of the following because of the risk of catching or passing on coronavirus at the moment?” (asked prior to seeing mock-up) and “If you had received the result you just saw, how worried would you feel doing each of the following because of the risk of catching or passing on coronavirus right now?” (asked after seeing mock-up): (Randomised presentation)  Shopping in a busy supermarket; Eating indoors in a restaurant with a small group of friends; Drinking in a pub garden with a small group of friends; Going to a large cinema; Travelling on the London underground; Visiting an elderly person in a nursing home; Attending Accident and Emergency in a city hospital  If the person who got this result caught COVID-19, how likely do you think it is that they would die as a result?  How would you describe the risk of this person dying from COVID-19 if they caught it? | Mean score on 13 4-point Likert scales (Q1-3 not at all satisfied – very satisfied; Q4-8 not easy at all – very easy; Q9-13 not effectively at all – very effectively)  Mean score on 5 7-point Likert scales (not at all – completely)  7-point Likert scale from 1 (“not at all”) to 7 (“completely”)  Free text  Free text    7-point Likert scales: Completely disagree - Completely agree  7-point Likert scales: Not at all worried - Very worried  7-point Likert scale: Not at all worried – Very worried  slider with no numerical cues: ‘very low risk’ and ‘very high risk’ as the end points  Above 2 combined into an index measure of cognitive risk perception (r=0.82) |
| Subscale 9, “Understanding health information well enough to know what to do,” of the Health Literacy Questionnaire [49] | 2, 3, 4 | It is important we understand how confident you are with reading and understanding health information. Please indicate how difficult or easy the following tasks are for you now. *Confidently fill medical forms in the correct way*  *Accurately follow the instructions from healthcare providers*  *Read and understand written health information*  *Read and understand all the information on medication labels*  *Understand what healthcare providers are asking you to do* | Labelled 5-point scale: Cannot do or always difficult (1); usually difficult (2); sometimes difficult (3); usually easy (4); always easy (5) |
| Numeracy questions  (the adaptive Berlin numeracy test [45], 3 items from [46] and a single item from [47]) | 1, 2, 3, 4 | Out of 1,000 people in a small town 500 are members of a choir. Out of these 500 members in the choir 100 are men.  Out of the 500 inhabitants that are not in the choir 300 are men. What is the probability that a randomly drawn man is a member of the choir?  Imagine we are throwing a five-sided die 50 times. On average, out of these 50 throws how many times would this five-sided die show an odd number (1, 3 or 5)?  Imagine we are throwing a loaded die (6 sides). The probability that the die shows a 6 is twice as high as the probability of each of the other numbers. On average, out of these 70 throws how  many times would the die show the number 6?  In a forest 20% of mushrooms are red, 50% brown and 30% white. A red mushroom is poisonous with a probability of 20%. A mushroom that is not red is poisonous with a probability of 5%. What is the probability that a poisonous mushroom in the forest is red?  Which of the following numbers represents the biggest risk of getting a disease?  1 in 100; 1 in 1000; 1 in 10  Imagine that we flip a fair coin 1,000 times. What is your best guess about how many times the coin would come up heads in 1,000 flips?  In a scratch card lottery, the chance of winning a £10 prize on the card is 1%. What is your best guess about how many people would win a £10 prize if 1,000 people each buy a single scratch card?  At a raffle, the chance of winning a car is 1 in 1,000. What percentage of tickets in the raffle win a car? |  |

### Appendix 4

Regression outputs for experiments in Surveys 2 and 4 looking at the format of the number (percentage v frequency) and contextual information (eg. age comparators, personas).

1) Estimates from regression predicting risk ratings (‘no information’ condition only)

|  | b | SE | β |
| --- | --- | --- | --- |
| (Intercept) | 45.76*** | 4.30 | 0.00 |
| Age^1^ | -0.76 | 0.53 | -0.04 |
| Sex (Male) | 1.04 | 1.68 | 0.02 |
| Risk level | 0.23 | 0.30 | 0.06 |
| Numeracy | -4.85*** | 0.66 | -0.31 |
| Format (Frequency) | -5.15*** | 1.20 | -0.18 |
| Risk level*Numeracy | 0.28*** | 0.06 | 0.38 |
| Format*Numeracy | 0.02 | 0.11 | 0.01 |
| Adjusted *R*^2^ | .233 |  |  |

^1^Age recorded as one of six brackets

****p* < .001

2) Estimates from exploratory regression predicting risk ratings (including all conditions)

|  | b | SE | β |
| --- | --- | --- | --- |
| (Intercept) | 43.44*** | 4.19 | 0.00 |
| Age^1^ | 0.03 | 0.30 | 0.00 |
| Sex (Male) | -1.55 | 0.97 | -0.02 |
| Risk level | 0.23 | 0.33 | 0.05 |
| Numeracy | -4.78*** | 0.79 | -0.29 |
| Group^2^ (Comparison information) | -1.57 | 4.99 | -0.02 |
| Group (Consonant context) | 6.91 | 4.95 | 0.09 |
| Group (Dissonant context) | 36.96*** | 4.85 | 0.49 |
| Format (Frequency) | 7.04** | 2.44 | 0.11 |
| Risk level*Numeracy | 0.28*** | 0.07 | 0.34 |
| Risk level*Group (Comparison information) | -0.05 | 0.47 | -0.01 |
| Risk level*Group (Consonant context) | 1.38** | 0.46 | 0.21 |
| Risk level*Group (Dissonant context) | -2.14*** | 0.45 | -0.33 |
| Numeracy*Group (Comparison information) | 0.62 | 1.01 | 0.04 |
| Numeracy*Group (Consonant context) | 1.20 | 1.01 | 0.08 |
| Numeracy*Group (Dissonant context) | -0.14 | 1.01 | -0.01 |
| Numeracy*Format | -0.25 | 0.50 | -0.02 |
| Risk level*Numeracy*Group (Comparison information) | 0.11 | 0.09 | 0.09 |
| Risk level*Numeracy*Group (Consonant context) | -0.12 | 0.09 | -0.09 |
| Risk level*Numeracy*Group (Dissonant context) | -0.04 | 0.09 | -0.03 |
| Adjusted *R*^2^ | .272 |  |  |

^1^Age recorded as one of six brackets. ^2^‘No information’ group is reference category

***p* < .01, ****p* < .001

Pairwise Levene’s tests for the experiment comparing different formats of absolute risk presentation

| Risk Level | Group 1 | Group 2 | Statistic |
| --- | --- | --- | --- |
| 0.1% | Percentage | X in 100 | *F*(1, 876)=8.92, *p* < .01 |
| 0.1% | Percentage | X in 1000 | *F*(1, 872)=32.58, *p* < .001 |
| 0.1% | Percentage | 1 in X | *F*(1, 890)=46.21, *p* < .001 |
| 0.1% | X in 100 | X in 1000 | *F*(1, 872)=7, *p* < .01 |
| 0.1% | X in 100 | 1 in X | *F*(1, 890)=10.94, *p* < .001 |
| 0.1% | X in 1000 | 1 in X | *F*(1, 886)=0.08, *p*=0.77 |
| 1% | Percentage | X in 100 | *F*(1, 876)=30.19, *p* < .001 |
| 1% | Percentage | X in 1000 | *F*(1, 872)=31.62, *p* < .001 |
| 1% | Percentage | 1 in X | *F*(1, 890)=67.94, *p* < .001 |
| 1% | X in 100 | X in 1000 | *F*(1, 872)=0.17, *p*=0.68 |
| 1% | X in 100 | 1 in X | *F*(1, 890)=3.11, *p*=0.08 |
| 1% | X in 1000 | 1 in X | *F*(1, 886)=6.31, *p* < .05 |
| 5% | Percentage | X in 100 | *F*(1, 876)=12.06, *p* < .001 |
| 5% | Percentage | X in 1000 | *F*(1, 872)=21.52, *p* < .001 |
| 5% | Percentage | 1 in X | *F*(1, 890)=24.65, *p* < .001 |
| 5% | X in 100 | X in 1000 | *F*(1, 872)=1.31, *p*=0.25 |
| 5% | X in 100 | 1 in X | *F*(1, 890)=2.6, *p*=0.11 |
| 5% | X in 1000 | 1 in X | *F*(1, 886)=0.29, *p*=0.59 |
| 12% | Percentage | X in 100 | *F*(1, 876)=0.97, *p*=0.33 |
| 12% | Percentage | X in 1000 | *F*(1, 872)=2.61, *p*=0.11 |
| 12% | Percentage | 1 in X | *F*(1, 890)=5.13, *p* < .05 |
| 12% | X in 100 | X in 1000 | *F*(1, 872)=0.42, *p*=0.52 |
| 12% | X in 100 | 1 in X | *F*(1, 890)=2.07, *p*=0.15 |
| 12% | X in 1000 | 1 in X | *F*(1, 886)=0.83, *p*=0.36 |
| 20% | Percentage | X in 100 | *F*(1, 876)=2.62, *p*=0.11 |
| 20% | Percentage | X in 1000 | *F*(1, 872)=0.26, *p*=0.61 |
| 20% | Percentage | 1 in X | *F*(1, 890)=0.35, *p*=0.55 |
| 20% | X in 100 | X in 1000 | *F*(1, 872)=1.58, *p*=0.21 |
| 20% | X in 100 | 1 in X | *F*(1, 890)=0.62, *p*=0.43 |
| 20% | X in 1000 | 1 in X | *F*(1, 886)=0.04, *p*=0.85 |

As an exploratory measure, we had also included a condition identical to the percentage condition, except respondents rated the risk on an 11pt Likert scale, not a slider, to see whether it made participants think less about what percentage along the slider they were moving their marker. A pre-registered comparison of estimates on this scale (rescaled) with those entered using a slider (the percentage condition only) using a mixed two-way ANOVA (2(scale format; between)x5(risk level; within) which revealed a significant interaction, *F*(4, 3556)= 5.10, *p*<0.001, *η*^2^_G_=0.01. Pairwise comparisons (Tukey’s HSD) at each risk level revealed that the mean risk ratings did not differ significantly between groups who entered their response via a sliding scale or a Likert scale, except in the case of the 20% risk level condition, where Likert responses were slightly higher than slider responses (*M*_diff=_4.13, *p*<0.05, Cohen’s *d*=-0.15).

### Appendix 5

Results of logistic regression models predicting the likelihood of correct responses to objective comprehension items from information format and risk level.

|  | Percentage item | | | Frequency item | | |
| --- | --- | --- | --- | --- | --- | --- |
| *Predictors* | *OR* | *CI* | *p* | *OR* | *CI* | *p* |
| (Intercept) | 0.46 | 0.31 – 0.67 | <0.001 | 1.82 | 1.27 – 2.65 | 0.001 |
| Format [Scale,  negative, age comparison] | 1.45 | 0.87 – 2.45 | 0.156 | 0.79 | 0.47 – 1.31 | 0.36 |
| Format [Scale,  negative, 'flu  comparison] | 1.79 | 1.07 – 3.04 | 0.028 | 0.81 | 0.48 – 1.35 | 0.419 |
| Format [Scale,  negative, no comparison] | 1.05 | 0.61 – 1.80 | 0.857 | 1.09 | 0.64 – 1.84 | 0.757 |
| Format [Scale, positive, no comparison] | 0.85 | 0.49 – 1.46 | 0.551 | 0.52 | 0.31 – 0.86 | 0.011 |
| Risk level [0.1%] | 1.8 | 1.07 – 3.04 | 0.027 | 0.79 | 0.47 – 1.31 | 0.361 |
| Risk level [2%] | 3.12 | 1.85 – 5.32 | <0.001 | 0.92 | 0.54 – 1.54 | 0.744 |
| Risk level [20%] | 3.53 | 2.10 – 6.01 | <0.001 | 0.71 | 0.43 – 1.19 | 0.195 |
| Format [Scale,  negative, age comparison]  * Risk level [0.1%] | 0.49 | 0.24 – 1.01 | 0.055 | 1.08 | 0.53 – 2.20 | 0.842 |
| Format [Scale,  negative, 'flu  comparison] *  Risk level [0.1%] | 0.47 | 0.22 – 0.96 | 0.039 | 1.2 | 0.58 – 2.47 | 0.626 |
| Format [Scale,  negative, no comparison]  * Risk level [0.1%] | 0.99 | 0.48 – 2.06 | 0.983 | 1.01 | 0.49 – 2.09 | 0.982 |
| Format [Scale,  positive, no comparison]  * Risk level [0.1%] | 0.94 | 0.44 – 1.97 | 0.86 | 1.25 | 0.61 – 2.56 | 0.543 |
| Format [Scale,  negative, age comparison]  * Risk level [2%] | 0.96 | 0.46 – 2.00 | 0.914 | 1.16 | 0.56 – 2.41 | 0.68 |
| Format [Scale,  negative, 'flu  comparison] *  Risk level [2%] | 0.81 | 0.39 – 1.68 | 0.564 | 1.11 | 0.54 – 2.30 | 0.771 |
| Format [Scale,  negative, no comparison]  * Risk level [2%] | 0.86 | 0.41 – 1.81 | 0.697 | 0.98 | 0.47 – 2.06 | 0.961 |
| Format [Scale,  positive, no comparison]  * Risk level [2%] | 0.96 | 0.45 – 2.01 | 0.904 | 1.41 | 0.69 – 2.90 | 0.351 |
| Format [Scale,  negative, age comparison]  * Risk level [20%] | 0.6 | 0.29 – 1.24 | 0.169 | 1.08 | 0.53 – 2.20 | 0.837 |
| Format [Scale,  negative, 'flu  comparison] *  Risk level [20%] | 0.49 | 0.24 – 1.01 | 0.052 | 0.97 | 0.47 – 1.98 | 0.932 |
| Format [Scale,  negative, no comparison]  * Risk level [20%] | 1.08 | 0.52 – 2.28 | 0.83 | 1.05 | 0.51 – 2.17 | 0.893 |
| Format [Scale,  positive, no comparison]  * Risk level [20%] | 0.94 | 0.44 – 1.98 | 0.867 | 2.11 | 1.03 – 4.34 | 0.041 |
| Observations | 2496 | | | 2500 | | |
| R^2^ Tjur | 0.058 | | | 0.01 | | |

Reference categories: format: Text only, negative, age comparison; Risk level: 0.01%

### Appendix 6

Experiment 4.3 exploratory regression outputs and statistics, ordinary least squares. Each column represents a separate regression with the column header as the dependent variable. The format *‘scale, negative, no comparison’* was treated as the reference level.

|  | Cognitive risk perception | Emotional risk perception | Concern about higher-risk behaviours | Increase in concern about higher-risk behaviours | Communication efficacy | Actionability | Subjective clarity | Subjective comprehension |
| --- | --- | --- | --- | --- | --- | --- | --- | --- |
| Predictors | std. Beta  (std. CI) | std. Beta  (std. CI) | std. Beta  (std. CI) | std. Beta  (std. CI) | std. Beta  (std. CI) | std. Beta  (std. CI) | std. Beta  (std. CI) | std. Beta  (std. CI) |
| Format:  Scale, negative, age comparison | 1.73 (-1.18 – 4.63) | -0.37 (-3.87 – 3.12) | -0.04 (-0.23 – 0.14) | -0.03 (-0.18 – 0.11) | -0.09* (-0.17 – -0.0) | -0.18 (-0.38 – 0.02) | -0.15 (-0.33 – 0.03) | -0.08 (-0.25 – 0.1) |
| Format:  Scale, negative, ‘flu comparison | 1.05 (-1.84 – 3.94) | 0.3 (-3.17 – 3.77) | -0.09 (-0.27 – 0.09) | -0.03 (-0.17 – 0.11) | -0.1* (-0.18 – -0.02) | -0.07 (-0.27 – 0.12) | -0.11 (-0.29 – 0.07) | -0.13 (-0.3 – 0.04) |
| Format:  Scale, positive, no comparison | 0.59 (-2.31 – 3.49) | -1.75 (-5.24 – 1.73) | -0.29** (-0.47 – -0.11) | -0.17* (-0.32 – -0.03) | -0.07 (-0.15 – 0.01) | 0.04 (-0.16 – 0.23) | -0.07 (-0.25 – 0.11) | -0.09 (-0.26 – 0.08) |
| Format:  Text, negative, age comparison | 4.38** (1.5 – 7.25) | 2.49 (-0.97 – 5.95) | 0.08 (-0.1 – 0.26) | 0.08 (-0.06 – 0.23) | -0.03 (-0.11 – 0.05) | -0.1 (-0.3 – 0.09) | 0.08 (-0.1 – 0.26) | 0.03 (-0.14 – 0.2) |
| Risk level | 1.3*** (1.19 – 1.41) | 1.49*** (1.36 – 1.62) | 0.03*** (0.03 – 0.04) | 0.04*** (0.04 – 0.05) | -0.01*** (-0.01 – -0.0) | -0.01*** (-0.02 – -0.01) | 0.0 (-0.01 – 0.01) | -0.0 (-0.01 – 0.0) |
| Sex: Male | 4.81*** (2.93 – 6.69) | 2.89* (0.64 – 5.15) | -0.16** (-0.27 – -0.04) | 0.01 (-0.09 – 0.1) | 0.01 (-0.04 – 0.06) | 0.14* (0.02 – 0.27) | -0.01 (-0.13 – 0.11) | -0.04 (-0.15 – 0.07) |
| Age | -0.12*** (-0.17 – -0.06) | -0.01 (-0.08 – 0.06) | 0.01*** (0.01 – 0.01) | 0.01*** (0.0 – 0.01) | 0.003*** (0.0 – 0.01) | 0.01** (0.0 – 0.01) | 0.0 (-0.0 – 0.01) | 0.0 (-0.0 – 0.0) |
| Numeracy | -4.71*** (-5.21 – -4.22) | -3.53*** (-4.13 – -2.93) | -0.06*** (-0.09 – -0.03) | 0.0 (-0.02 – 0.03) | 0.03*** (0.02 – 0.05) | -0.04* (-0.07 – -0.01) | 0.12*** (0.08 – 0.15) | 0.17*** (0.14 – 0.2) |
| Prior COVID-19 risk perception | 2.74*** (1.8 – 3.68) | 6.96*** (5.83 – 8.08) | 0.64*** (0.59 – 0.7) | -0.14*** (-0.18 – -0.09) | 0.03* (0.0 – 0.05) | -0.03 (-0.1 – 0.03) | 0.02 (-0.04 – 0.07) | 0.03 (-0.03 – 0.08) |
| Observations (N) R^2^ / R^2^ adjusted | 2184 .306/.304 | 2188 .262/.259 | 2189 .228/.225 | 2189 .114/.110 | 2189 .028/.024 | 2186 .018/.014 | 2189 .028/.024 | 2188 .059/.055 |

Note: * p<0.05   ** p<0.01   *** p<0.001

Experiment 4.3 regression outputs and statistics, ordinary least squares (contd.)

|  | Agreement with "If this result applied to me, I would not be worried about catching COVID-19" | Agreement with "If this result applied to me, I would likely be anxious and it might affect my mental health" | Agreement with "If this result applied to me, I would likely change my behaviour to be less cautious of catching the virus" | Agreement with "If this result applied to me, I would do everything I could to avoid catching the virus" | Agreement with “If this result applied to me I'd be more concerned about my risk from COVID-19 than from seasonal 'flu” |
| --- | --- | --- | --- | --- | --- |
| Predictors | std. Beta  (std. CI) | std. Beta  (std. CI) | std. Beta  (std. CI) | std. Beta  (std. CI) | std. Beta  (std. CI) |
| Format:  Scale, negative, age comparison | 0.09 (-0.16 – 0.33) | -0.15 (-0.37 – 0.08) | -0.08 (-0.34 – 0.18) | 0.00 (-0.21 – 0.21) | 0.03 (-0.20 – 0.27) |
| Format:  Scale, negative, ‘flu comparison | 0.06 (-0.18 – 0.3) | 0.06 (-0.17 – 0.28) | -0.17 (-0.43 – 0.09) | 0.04 (-0.16 – 0.25) | 0.21 (-0.02 – 0.44) |
| Format:  Scale, positive, no comparison | 0.39** (0.15 – 0.64) | -0.36** (-0.58 – -0.13) | -0.03 (-0.29 – 0.22) | -0.17 (-0.38 – 0.03) | -0.25* (-0.49 – -0.02) |
| Format:  Text, negative, age comparison | -0.14 (-0.38 – 0.1) | 0.15 (-0.07 – 0.38) | -0.05 (-0.30 – 0.21) | 0.08 (-0.13 – 0.29) | 0.18 (-0.04 – 0.41) |
| Risk level | -0.06*** (-0.07 – -0.05) | 0.06*** (0.06 – 0.07) | 0.02** (0.01 – 0.03) | 0.04*** (0.03 – 0.05) | 0.07*** (0.06 – 0.08) |
| Sex: Male | 0.23** (0.07 – 0.39) | 0.18* (0.04 – 0.33) | 0.22* (0.05 – 0.38) | -0.21** (-0.34 – -0.07) | 0.10 (-0.04 – 0.26) |
| Age | -0.01*** (-0.01 – -0.01) | -0.02*** (-0.02 – -0.01) | -0.02*** (-0.02 – -0.01) | 0.02*** (0.01 – 0.02) | 0.0 (0.00 – 0.01) |
| Numeracy | -0.11*** (-0.15 – -0.06) | -0.26*** (-0.30 – -0.22) | -0.25*** (-0.3 – -0.21) | -0.08*** (-0.11 – -0.04) | -0.13*** (-0.17 – 0.09) |
| Prior COVID-19 risk perception | -0.39*** (-0.47 – -0.31) | 0.48*** (0.40 – 0.55) | 0.08 (-0.00 – 0.16) | 0.46*** (0.39 – 0.52) | 0.47*** (0.4 – 0.55) |
| Observations (N) R^2^ / R^2^ adjusted | 2189 .131/.127 | 2189 .221/.218 | 2189 .083/.079 | 2189 .153/.149 |  |

### Appendix 7

Experiment 4.3 exploratory logistic regression outputs and statistics. Each column represents a separate regression with the column header as the dependent variable. The format *‘scale, negative, no comparison’* was treated as the reference level.

|  | Objective comprehension (percentages) | Objective comprehension (frequencies) |
| --- | --- | --- |
| Predictors | OR  (std. CI) | OR  (std. CI) |
| Format:  Scale, negative, age comparison | 1.28 (0.95 – 1.73) | 0.81 (0.59 – 1.11) |
| Format:  Scale, negative, ‘flu comparison | 1.28 (0.95 – 1.73) | 0.72* (0.53 – 0.98) |
| Format:  Scale, positive, no comparison | 0.74* (0.55 – 0.99) | 0.50*** (0.37 – 0.68) |
| Format:  Text, negative, age comparison | 0.97 (0.72 – 1.31) | 0.79 (0.58 – 1.08) |
| Risk level | 1.05*** (1.03 – 1.06) | 0.99 (0.98 – 1.0) |
| Sex: Male | 0.73** (0.6 – 0.9) | 0.62*** (0.5 – 0.76) |
| Age | 1.01* (1.0 – 1.01) | 1.01** (1.01 – 1.02) |
| Numeracy | 1.77*** (1.67 – 1.9) | 1.79*** (1.68 – 1.9) |
| Prior COVID-19 risk perception | 0.99 (0.9 – 1.08) | 1.12* (1.01 – 1.23) |
| Observations (N) Pseudo-R^2^ | 2188  .166 | 2189  .164 |
